# Exclusion of bacterial co-infection in COVID-19 using baseline inflammatory markers and their response to antibiotics

**DOI:** 10.1101/2020.10.09.20199778

**Authors:** Claire Y Mason, Tanmay Kanitkar, Charlotte J Richardson, Marisa Lanzman, Zak Stone, Tabitha Mahungu, Damien Mack, Emmanuel Q Wey, Lucy Lamb, Indran Balakrishnan, Gabriele Pollara

## Abstract

**Background:** COVID-19 is infrequently complicated by secondary bacterial infection, but nevertheless antibiotic prescriptions are common. We used community-acquired pneumonia (CAP) as a benchmark to define the processes that occur in a bacterial pulmonary infection, and tested the hypothesis that baseline inflammatory markers and their response to antibiotic therapy could distinguish CAP from COVID-19.

**Methods:** In patients admitted to Royal Free Hospital (RFH) and Barnet Hospital (BH) we defined CAP by lobar consolidation on chest radiograph, and COVID-19 by SARS-CoV-2 detection by PCR. Data were derived from routine laboratory investigations.

**Results:** On admission all CAP and >90% COVID-19 patients received antibiotics. We identified 106 CAP and 619 COVID-19 patients at RFH. CAP was characterised by elevated white cell count (WCC) and C-reactive protein (CRP) compared to COVID-19 (median WCC 12.48 (IQR 8.2-15.3) vs 6.78 (IQR 5.2-9.5) x10^6^ cells/ml and median CRP CRP 133.5 (IQR 65-221) vs 86 (IQR 42-160) mg/L). Blood samples collected 48-72 hours into admission revealed decreasing CRP in CAP but not COVID-19 (CRP difference −33 (IQR −112 to +3.5) vs +15 (IQR −15 to +70) mg/L respectively). In the independent validation cohort (BH) consisting of 169 CAP and 181 COVID-19 patients, admission WCC >8.2×10^6^ cells/ml or falling CRP during admission identified 95% of CAP cases, and predicted the absence of bacterial co-infection in 45% of COVID-19 patients.

**Conclusions:** We propose that in COVID-19 the absence of both elevated baseline WCC and antibiotic-related decrease in CRP can exclude bacterial co-infection and facilitate antibiotic stewardship efforts.

## Introduction

The COVID-19 pandemic caused by the novel beta coronavirus SARS-CoV-2 has caused over 34 million infections and over 1 million deaths worldwide [1]. The drivers of pathology remain to be elucidated, but a hyperinflammatory response is associated with worse case fatality [2]. Other viral respiratory tract infections, best characterised by influenza, can be complicated by bacterial co-infections that also raise inflammatory markers and are associated with high mortality [3,4], but distinguishing severe viral pneumonia from bacterial co-infection is challenging [5]. In COVID-19, several studies have found bacterial co-infection to be rare, as determined by identification of causative pathogens [6–9]. However, routine microbiological culture takes several days, lacks sensitivity [10] and does not readily distinguish bacterial colonisation from infection. Moreover, microbiological respiratory tract sampling is not routinely performed in patients admitted with COVID-19 [9]. Therefore, despite guidance aimed at rationalising antibiotic use [11], it is unsurprising that diverse and elevated rates of antibiotic prescriptions have been reported in patients admitted for COVID-19 infection [8,9].

It is likely that many COVID-19 associated antibiotic prescriptions are given in the absence of bacterial co-infection, thus hampering antimicrobial stewardship efforts and potentially increasing antimicrobial resistance [12–14]. Many studies have focused on clinical and laboratory features that risk stratify outcome in COVID-19 [15–18], but currently infections caused by virus alone cannot be readily distinguished from those with a bacterial component. C-reactive protein (CRP), white cell count (WCC) and procalcitonin (PCT) have been used to distinguish between influenza and bacterial pneumonia, allowing antibiotic treatment to be omitted or stopped [19–22]. Serial measurements of inflammatory markers may also assist in distinguishing bacterial from viral infections [23,24]. A small retrospective study comparing COVID-19 to community-acquired pneumonia patients identified differences in admission neutrophil counts, D-dimers and CRP, but did not provide a rigorous definition for the pneumonia cases, nor explored changes in these markers over time [25].

In this study we aimed to identify features that discriminated viral COVID-19 infections from those complicated by bacterial co-infection. We used community-acquired pneumonia (CAP) as a benchmark to define the processes that occur in bacterial pulmonary infections, and tested the hypothesis that baseline inflammatory markers and their response to antibiotics could distinguish CAP from most COVID-19 infections. To address this research question, we performed a retrospective, cohort study from a large split-site academic hospital in the UK. We used the independent nature of the two sites to discover and validate our findings, extending their generalisability.

## Methods

### Data extraction and ethics

Anonymised demographics, antimicrobial prescriptions, haematological and biochemical investigations were extracted from the Clinical Practice Group analysis team, Cerner Electronic Patient Records and the electronic Clinical Infection Database (elCID), and microbiological investigations from WinPath at Royal Free London (RFL) NHS Trust [26]. The study was approved by the Research and Innovation Group at RFL NHS Trust, which stated that confidential patient information could be used under COVID-19 COPI notice made by Department of Health and Social Care, and that as this was a retrospective review of routine clinical data, formal ethical approval was not required.

### Patient selection

We identified patients from 2 hospital sites of RFL NHS Trust in London, UK: Royal Free Hospital (RFH) and Barnet Hospital (BH). These hospitals are separated by 11 kilometres and patient care is delivered by non-overlapping clinical staff, using non-identical clinical care bundles and antibiotic policies. We included patients aged >18 years old admitted to hospital, of which a subset was admitted for >48 hours (tables 1 & 4), and excluded patients with haematological malignancies. We defined COVID-19 and influenza patients by RT-PCR detection of SARS-CoV-2 and influenza A or B viruses respectively from nasopharyngeal swabs. COVID-19 patients were identified between 1^st^ March and 31^st^ May 2020, and influenza patients between 1^st^ January and 31^st^ May 2019. These criteria yielded 619 and 181 COVID-19 patients from RFH and BH respectively, and 188 and 162 influenza patients from RFH and BH respectively. There were no patients co-infected with SARS-CoV-2 and influenza viruses. CAP patients were identified by a clinical diagnosis of CAP made between 1^st^ January and 31^st^ May 2019 with focal consolidation on chest radiograph reported by consultant radiologists (106 patients at RFH and 169 at BH).

**Table 1.**
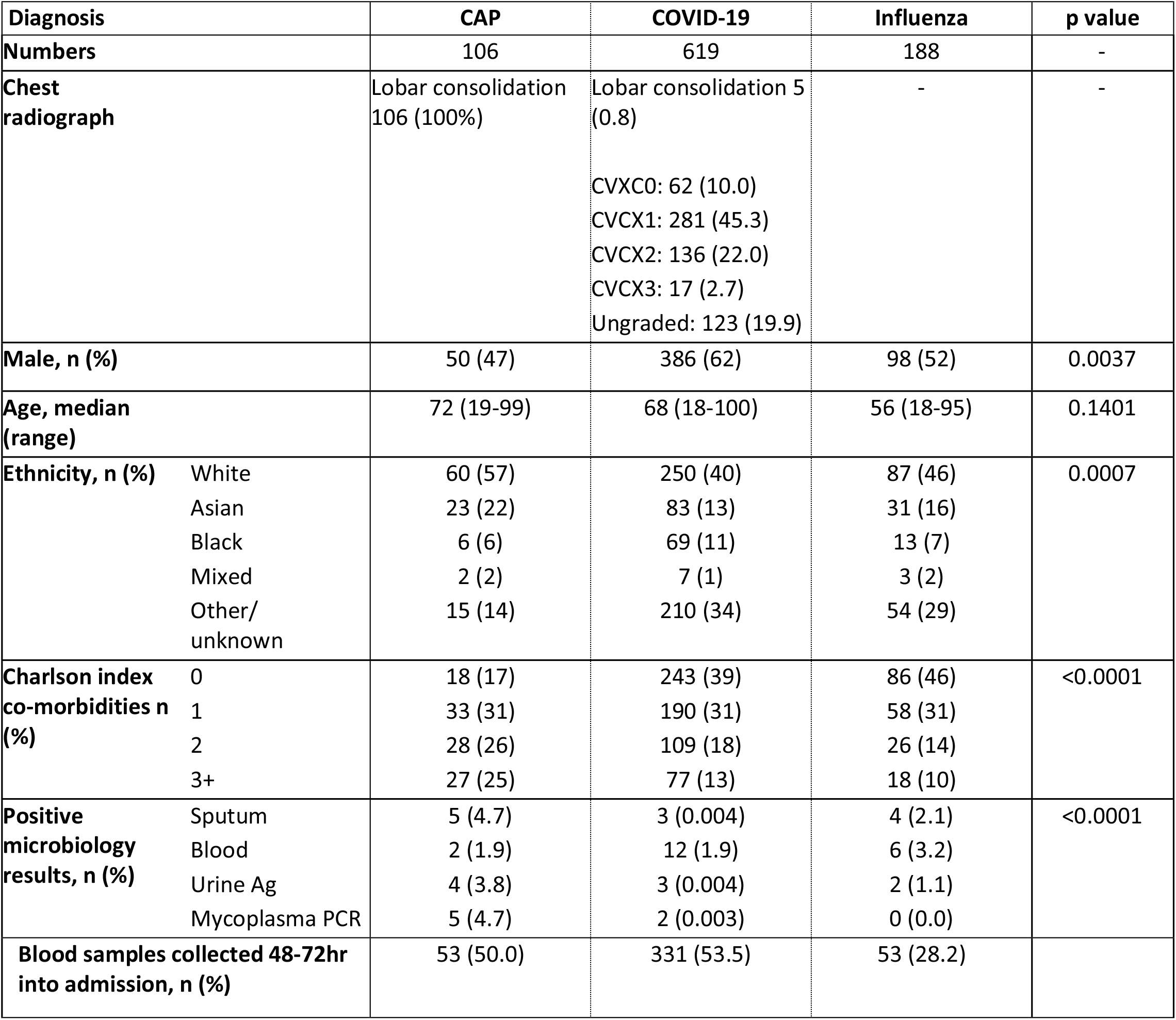
Patients identified in each diagnostic group at RFH. Chest radiograph codes for COVID-19 patients based on British Society of Thoracic Imaging guidelines, CVCX0 = Normal, CVCX1 = Classic for COVID-19, CVCX2 = Indeterminate for COVID-19, CVCX3 = Non-COVID-19. p values represent comparisons between CAP and COVID-19.

For some analyses, we excluded COVID-19 patients with radiological or microbiological evidence of ongoing bacterial co-infection. This was defined by presence of lobar pneumonia on a chest radiograph within 72 hours of hospital admission, or by a non-contaminant bacterial growth on blood culture, growth in sputum samples of *Streptococcus pneumoniae, Staphylococcus aureus* or *Moraxella catarrhalis*, detection of *Mycoplasma pneumoniae* by PCR from sputum or detection of *Streptococcus pneumoniae* antigen in urine (tables 1 & 3 and table S1).

### Statistical analysis

Baseline demographics were compared by Mann-Whitney test (age), Fisher’s exact test (gender) or Chi-square test (ethnicity, Charlson co-morbidities and microbiological results). Continuous variables from blood tests were expressed as median and interquartile range (IQR), and patient groups were compared using the non-parametric two-tailed Mann-Whitney U test. A multivariate logistic regression model was used to determine factors that discriminated between CAP and COVID-19. The diagnosis of CAP was the categorical output variable, and blood tests were used as continuous dependent variables in the model, which generated Receiver Operating Characteristic curves (ROC) and areas under the curve (AUC) as a summary statistic. For pre-determined cut-offs, we also calculated sensitivity, specificity, positive and negative predictive values, and positive and negative likelihood ratios. All analyses were performed using Microsoft Excel and GraphPad Prism version 8.

## Results

### Defining the discovery cohort

We identified 107 CAP, 620 COVID-19 and 188 influenza patients at RFH. 2 patients were excluded due to haematological malignancy. Male gender was overrepresented in COVID-19 (62% in COVID-19 vs 47% and 52% in CAP and influenza respectively), whereas CAP patients were older (median age 72, 68 and 56 in CAP, COVID-19 and influenza respectively). The proportion of Black, Asian, Mixed and Other (non-white) ethnicity patients was higher in COVID-19 compared to CAP and patients with CAP had more comorbidities and identified bacteria in routine microbiological investigations more commonly (table 1).

### Distinguishing pneumonia from COVID-19

We tested the hypothesis that inflammatory markers could discriminate CAP from COVID-19 or influenza by comparing total WCC, its differential cell counts and CRP levels on the day of admission to hospital. Compared to CAP, COVID-19 was associated with significantly lower median WCC (12.48 vs 6.78×10^6^/ml) and neutrophils (9.98 vs 5.37×10^6^/ml (fig 1)). Influenza was also characterised by significantly lower median WCC (7.28×10^6^/ml) and neutrophils (median 5.10×10^6^/ml) than pneumonia (fig 1). Lymphocyte counts did not differ between the groups. CRP was also significantly higher in CAP than in COVID-19, and lower in influenza (median CRP 133.5, 86.0 and 31mg/L respectively) (fig 1).

**Figure 1.**
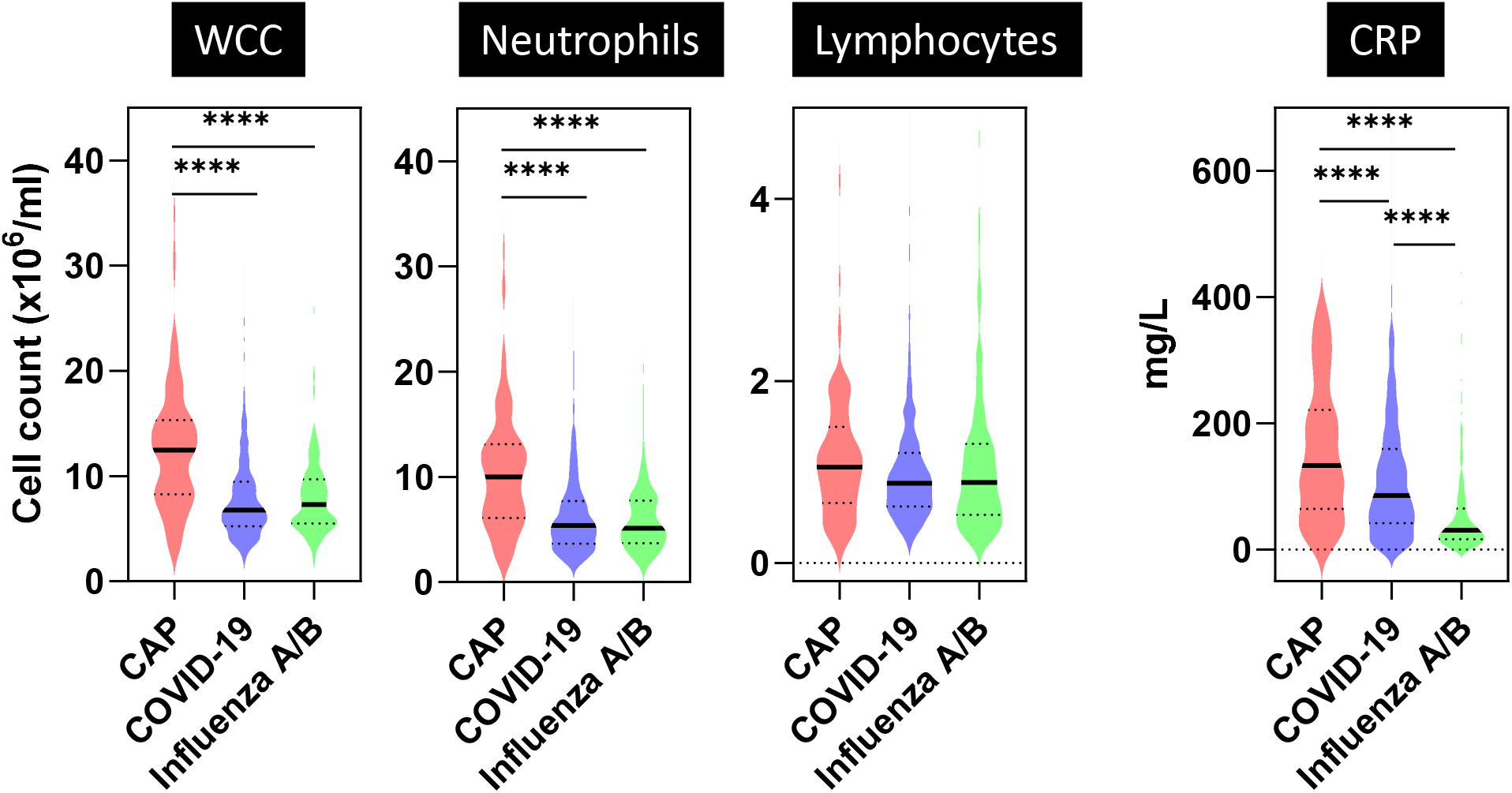
Admission blood samples for all patients admitted to RFH. Violin plots represent distribution of values for CAP (n=106), COVID-19 (n=619) and influenza A/B (n=188) patients. Bold lines represent median values. Dotted lines represent IQR values. **** indicates p<0.0001 by Mann-Whitney test.

All CAP patients were prescribed antibiotics on admission and in two independent surveys of COVID-19 patients from RFH, 95/100 (95%) and 104/118 (88%) were prescribed antibiotics to treat a presumptive superadded pulmonary bacterial infection. We hypothesised that CAP and viral infections could be further discriminated by changes in inflammatory markers following initiation of antibiotics [23]. In the RFH cohort, 53 (50%) CAP, 331 (53%) COVID-19 and 53 (28%) influenza patients were admitted for >48 hours and had a blood sample collected as part of routine clinical care 48-72 hours into admission (table 1). Differences in inflammatory markers on admission within this subset mirrored that seen in the wider cohort (fig S1). At this later time point, CAP was still characterised by elevated median WCC (9.5 vs 7.01×10^6^/ml), but this difference was diminished compared to admission (fig S2). Moreover, the difference in CRP between CAP and COVID-19 was no longer evident (median CRP 107.5 vs 127.0mg/L) (fig S2). These changes were driven by a greater fall in WCC and CRP for CAP compared to COVID-19 (ΔWCC −2.32 vs −0.17×10^6^/ml and ΔCRP −33 vs +15mg/L respectively) (fig 2).

**Figure 2.**
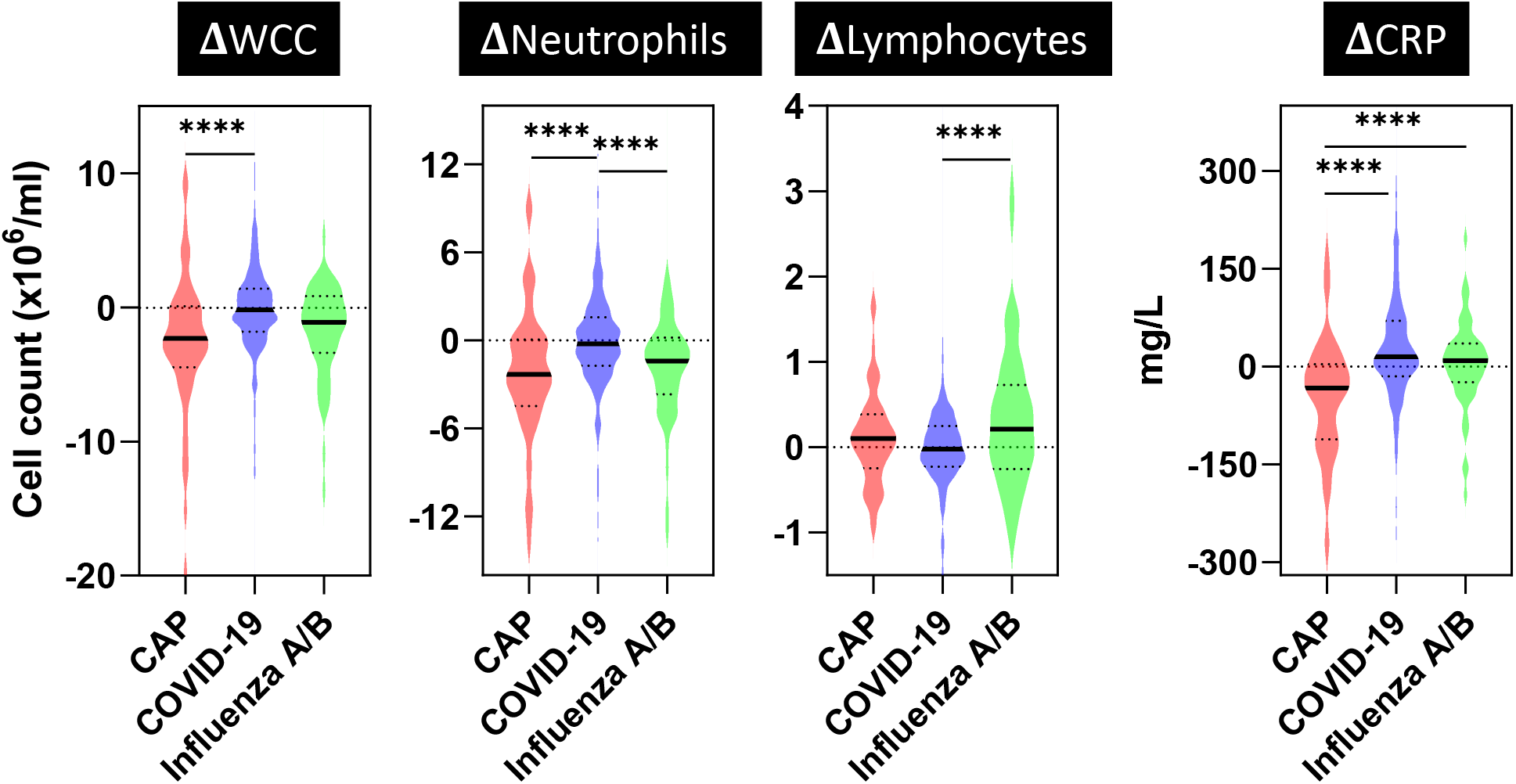
Change in values between admission blood samples and those collected 48-72 hours into admission at RFH. Violin plots represent distribution of difference (⍰) in investigation results between those collected on hospital admission and 48-72 hours into admission in CAP (n=53), COVID-19 (n=331) and influenza A/B (n=53) patients. Bold lines represent median values. Dotted lines represent IQR values. **** indicates p<0.0001 by Mann-Whitney test.

### Contribution of multiple variables to discriminate pneumonia from COVID-19

Our data suggested that elevated WCC and CRP, as well as a reduction in these parameters at 48-72 hours could discriminate between COVID-19 and CAP. To test this hypothesis, we applied a logistic regression model to the data collected from these two patient groups. We used the diagnosis of CAP as the binary outcome variable and observed that WCC or ΔCRP measurements alone yielded the greatest diagnostic accuracy (fig 3 & table 2). The maximal AUC obtained from this analysis was 0.75 (C.I. 0.68-0.83), demonstrating the trade off in sensitivity and specificity when using these variables alone. We also tested whether their combined use would improve the diagnostic accuracy of the model, and observed some improvement in discriminating between CAP from COVID-19 (AUC 0.80, C.I. 0.73-0.87), with no added benefit to also including ΔWCC measurements (fig 3 & table 2).

**Table 2.**
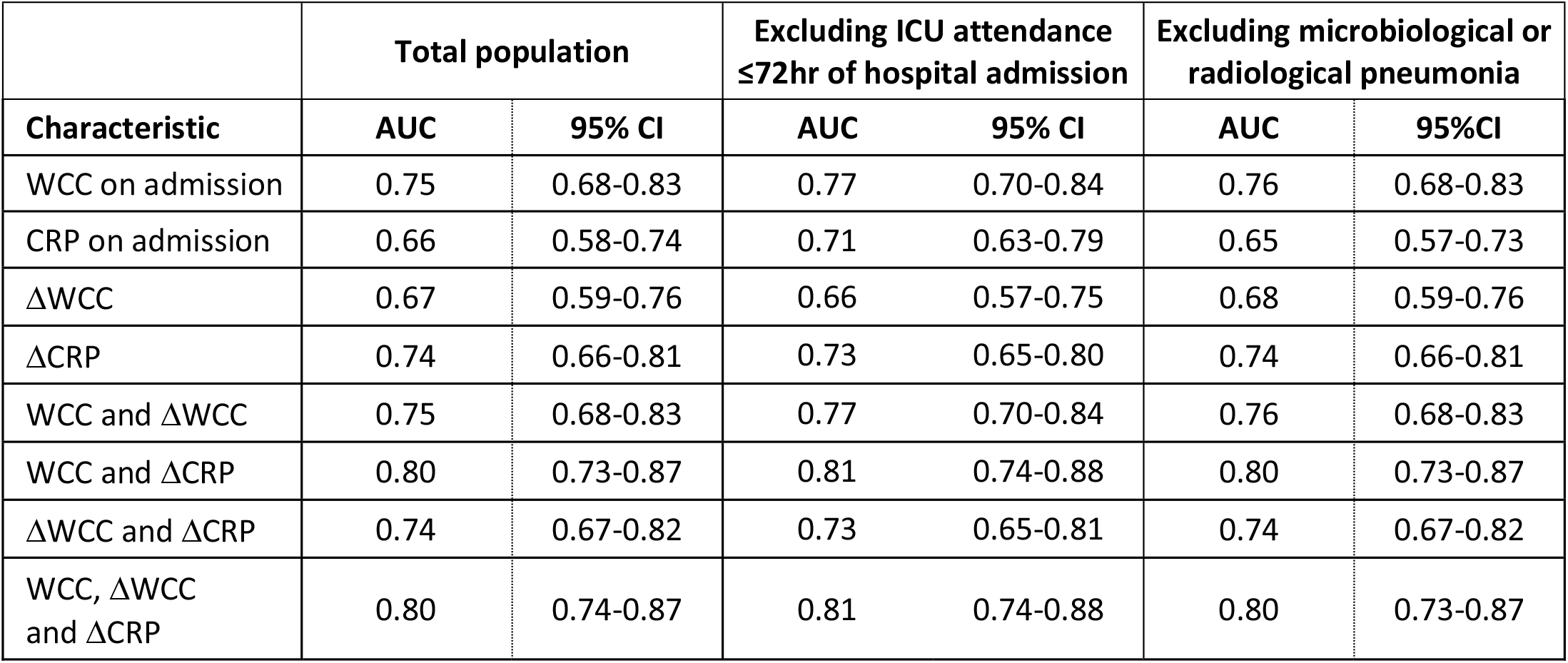
Discriminatory accuracy of WCC, CRP, ΔWCC and ΔCRP for diagnosis of CAP compared to COVID-19 in RFH patients. Populations included were all patients (n=384) admitted >48 hours (n=53 for CAP and n=331 for COVID 19) or excluding 79 COVID-19 patients that attended ICU within 72 hours of hospital admission, or excluding 18 COVID-19 patients with microbiological or radiological evidence of pneumonia. AUC, area under the curve; CI, confidence interval.

|  | Total population |  | Excluding ICU attendance<br>≤72hr of hospital admission |  | Excluding microbiological or<br>radiological pneumonia |  |
| --- | --- | --- | --- | --- | --- | --- |
| Characteristic | AUC | 95% CI | AUC | 95% CI | AUC | 95%CI |
| WCC on admission | 0.75 | 0.68-0.83 | 0.77 | 0.70-0.84 | 0.76 | 0.68-0.83 |
| CRP on admission | 0.66 | 0.58-0.74 | 0.71 | 0.63-0.79 | 0.65 | 0.57-0.73 |
| ΔWCC | 0.67 | 0.59-0.76 | 0.66 | 0.57-0.75 | 0.68 | 0.59-0.76 |
| ΔCRP | 0.74 | 0.66-0.81 | 0.73 | 0.65-0.80 | 0.74 | 0.66-0.81 |
| WCC and ΔWCC | 0.75 | 0.68-0.83 | 0.77 | 0.70-0.84 | 0.76 | 0.68-0.83 |
| WCC and ΔCRP | 0.80 | 0.73-0.87 | 0.81 | 0.74-0.88 | 0.80 | 0.73-0.87 |
| ΔWCC and ΔCRP | 0.74 | 0.67-0.82 | 0.73 | 0.65-0.81 | 0.74 | 0.67-0.82 |
| WCC, ΔWCC<br>and ΔCRP | 0.80 | 0.74-0.87 | 0.81 | 0.74-0.88 | 0.80 | 0.73-0.87 |

**Figure 3.**
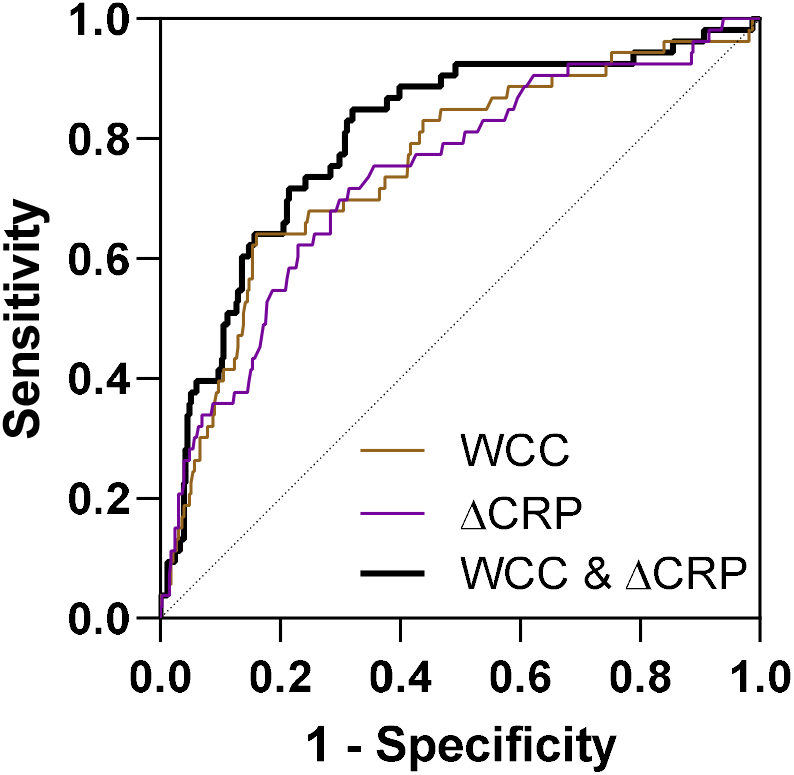
Accuracy of blood parameters to diagnose CAP in RFH patients. Receiver operating curves generated from logistic regression models that incorporate combinations of WCC on admission and difference (Δ) in CRP between samples on admission and 48-72 hours into admission in order to discriminate RFH patients diagnosed with CAP from those diagnosed with COVID-19.

We considered whether the ability of WCC and ΔCRP to discriminate between COVID-19 and CAP was confounded by admission to intensive care unit (ICU). At RFH, 79/331 (23.9%) of COVID-19 patients included in the logistic regression model were admitted to ICU within 72 hours of their admission (table 1). Excluding this subset of patients did not affect the discriminatory power of the model (AUC 0.81, C.I. 0.74-0.88) (table 2). We also considered whether COVID-19 patients with microbiological or radiological evidence of a bacterial infection could have confounded discrimination from pneumonia. We identified 16 (4.8%) COVID-19 patients included in the logistic regression model with microbiological results consistent with an active bacterial process (sputum culture positivity with a respiratory pathogen or a non-contaminant bacterial growth in blood cultures (table 1)), and 2 (0.1%) further COVID-19 patients with lobar pneumonia on chest radiograph (table 1). Excluding these 18 patients did not alter the differences observed in admission WCC or ΔCRP between CAP and COVID-19 (data not shown), and did not affect the classification accuracy of the remaining patients in the logistic regression model (table 2).

### Decision making criteria to discriminate bacterial pneumonia and COVID-19

We sought to convert our observations into practical decision-making criteria for clinical practice. We generated a series of cut-offs in the variables with greatest discriminatory power between CAP and COVID-19 (admission WCC and ΔCRP), and explored the trade off in sensitivity and specificity generated by these alone or in combination (table 3). For cut offs of WCC we used the lower quartile value of the CAP cohort (>8.2×10^6^/ml) and the upper quartile value of the COVID-19 cohort (>9.5×10^6^/ml). For cut offs of ΔCRP we used the lower quartile of the COVID-19 cohort (<-15mg/l) and the upper quartile of the CAP cohort (<3.5mg/l), rounded to 0mg/L for simplicity This revealed that using CAP-derived quartile cut-offs yielded greater sensitivity, at the expense of specificity. The lower prevalence of CAP in this cohort compared to COVID-19 offered a high negative predictive value (>90%). Requiring both a WCC>8.2×10^6^/ml and ΔCRP<0 improved specificity, at the expense of sensitivity. However, as such a strategy would result in many cases of CAP being missed, we also explored using the presence of either parameter to define CAP, yielding a sensitivity of >90%. Although the specificity of this approach was only 43%, the absence of both admission WCC>8.2×10^6^/ml and ΔCRP<0 could still exclude CAP, and by extension bacterial co-infection alongside COVID-19, promoting antibiotic cessation in 142 / 331 (43%) COVID-19 patients from this cohort (table 3).

**Table 3.** Discriminatory performance of WCC and ΔCRP cut-offs for diagnosis of bacterial pneumonia. Populations included were 366 patients admitted to hospital >48hours, including 52 pneumonia cases and 331 COVID-19 cases with no microbiological or radiological evidence of pneumonia.

| Cut-off | Sensitivity % | Specificity % | Positive predictive value % | Negative predictive value % | Positive likelihood ratio | Negative likelihood ratio |
| --- | --- | --- | --- | --- | --- | --- |
| WCC>8.2 | 79.2 | 58.8 | 24.6 | 94.4 | 1.92 | 0.35 |
| WCC>9.5 | 67.9 | 70.6 | 28.1 | 92.9 | 2.31 | 0.45 |
| ΔCRP<3.5 | 75.5 | 61.3 | 24.8 | 93.7 | 1.95 | 0.40 |
| ΔCRP<-15 | 62.3 | 75.0 | 29.7 | 92.2 | 2.50 | 0.50 |
| ΔCRP<0 | 73.6 | 65.2 | 26.4 | 93.6 | 2.11 | 0.41 |
| WCC>8.2 AND ΔCRP<0 | 62.3 | 80.8 | 35.4 | 92.6 | 3.25 | 0.47 |
| WCC>8.2 OR ΔCRP<0 | 90.6 | 43.1 | 21.2 | 96.4 | 1.59 | 0.22 |

### Independent cohort validation

To demonstrate reproducibility of our findings, we used independent patient cohorts from a separate hospital, BH, consisting of 169 CAP, 181 COVID-19 and 162 influenza A/B patients. To ensure comparability to the RFH cohort, the patients were identified over the same time periods using identical criteria. Baseline demographic analyses were comparable to those in the RFH cohort (table 4), and 99 (59%), 60 (33%) and 47 (29%) of CAP, COVID-19 and influenza patients respectively were admitted for >48 hours and had a blood sample collected as part of routine clinical care 48-72 hours into admission (table 4).

**Table 4.**
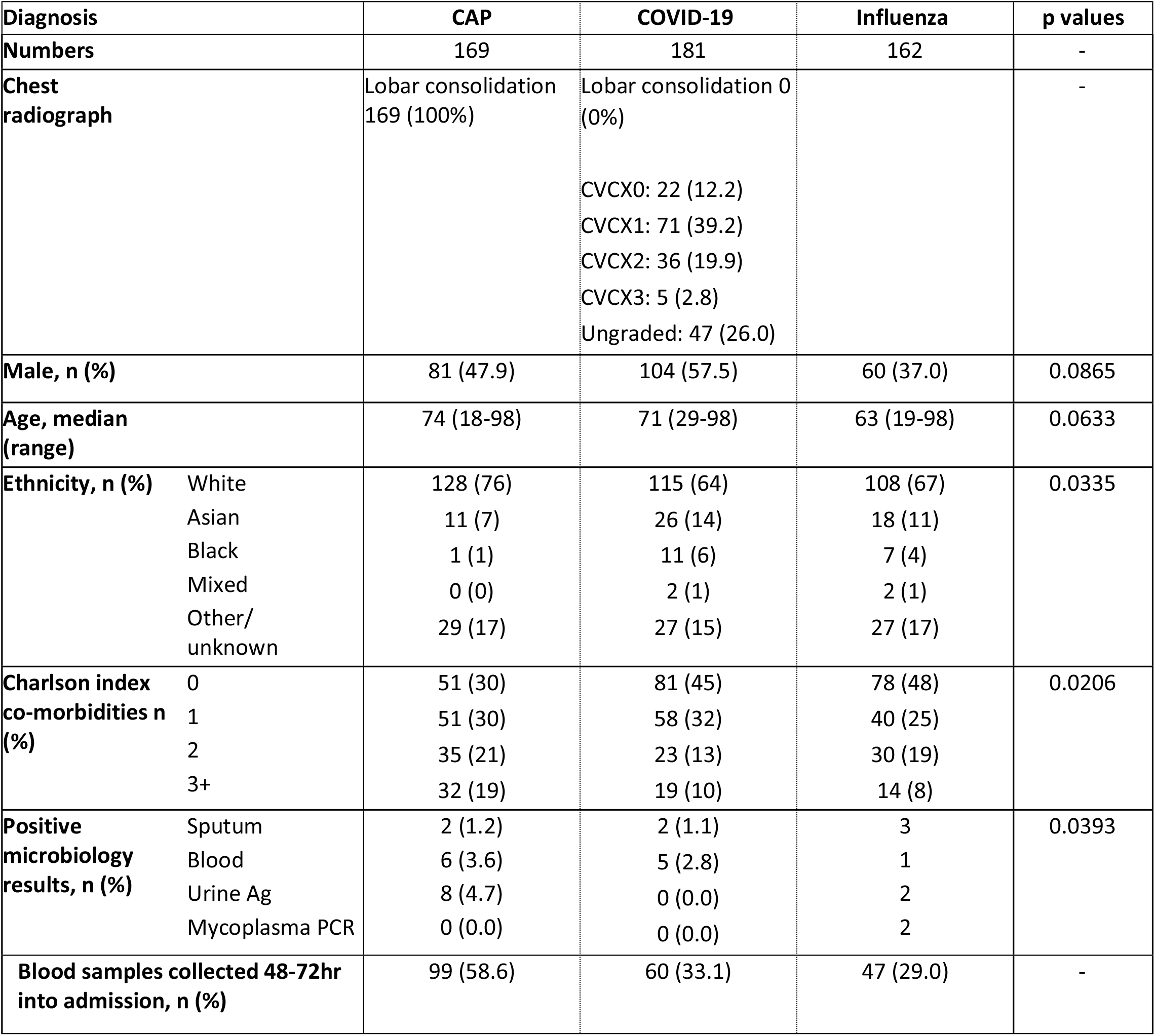
Patients identified in each diagnostic group at BH. Chest radiograph codes for COVID-19 patients based on British Society of Thoracic Imaging guidelines, CVCX0 = Normal, CVCX1 = Classic for COVID-19, CVCX2 = Indeterminate for COVID-19, CVCX3 = Non-COVID-19. p values represent comparisons between CAP and COVID-19.

| Diagnosis |  | CAP | COVID-19 | Influenza | p values |
| --- | --- | --- | --- | --- | --- |
| Numbers |  | 169 | 181 | 162 | - |
| Chest radiograph |  | Lobar consolidation<br>169 (100%) | Lobar consolidation 0<br>(0%)<br>CVCX0: 22 (12.2)<br>CVCX1: 71 (39.2)<br>CVCX2: 36 (19.9)<br>CVCX3: 5 (2.8)<br>Ungraded: 47 (26.0) |  | - |
| Male, n (%) |  | 81 (47.9) | 104 (57.5) | 60 (37.0) | 0.0865 |
| Age, median (range) |  | 74 (18-98) | 71 (29-98) | 63 (19-98) | 0.0633 |
| Ethnicity, n (%) | White | 128 (76) | 115 (64) | 108 (67) | 0.0335 |
|  | Asian | 11 (7) | 26 (14) | 18 (11) |  |
|  | Black | 1 (1) | 11 (6) | 7 (4) |  |
|  | Mixed | 0 (0) | 2 (1) | 2 (1) |  |
|  | Other/<br>unknown | 29 (17) | 27 (15) | 27 (17) |  |
| Charlson index co-morbidities n (%) | 0 | 51 (30) | 81 (45) | 78 (48) | 0.0206 |
|  | 1 | 51 (30) | 58 (32) | 40 (25) |  |
|  | 2 | 35 (21) | 23 (13) | 30 (19) |  |
|  | 3+ | 32 (19) | 19 (10) | 14 (8) |  |
| Positive microbiology results, n (%) | Sputum | 2 (1.2) | 2 (1.1) | 3 | 0.0393 |
|  | Blood | 6 (3.6) | 5 (2.8) | 1 |  |
|  | Urine Ag | 8 (4.7) | 0 (0.0) | 2 |  |
|  | Mycoplasma PCR | 0 (0.0) | 0 (0.0) | 2 |  |
| Blood samples collected 48-72hr into admission, n (%) |  | 99 (58.6) | 60 (33.1) | 47 (29.0) | - |

Differences in inflammatory markers within the BH cohort reflected those observed at RFH, with admission WCC and CRP levels being higher in CAP compared to COVID-19 and accompanied by a reduction following 48-72 hours of admission not observed in COVID-19 (fig S3). Applying these parameters into a logistic regression model demonstrated similar discriminatory power of each variable to that seen in RFH patients, and once more an optimal AUC (0.84, C.I. 0.78-0.91) was derived when both admission WCC and ΔCRP were included as variables in the model (table 5). We explored the exclusion of COVID-19 patients included in the logistic regression model that either attended ICU within 72 hours of hospital admission (21/60, 35.0%) or demonstrated microbiological or radiological evidence pneumonia (4/60, 6.7%) (table 5). However, as seen in the RFH cohort, excluding either subset of patients did not significantly affect the discriminatory ability of the logistic regression model (table 5).

**Table 5.** Discriminatory accuracy of WCC, CRP, ΔWCC and ΔCRP for detection of bacterial pneumonia in BH patients. Populations included were all patients (n=159) admitted >48 hours (n=99 for CAP and n=60 for COVID 19) or excluding 21 COVID-19 patients that attended ICU within 72 hours of hospital admission, or 4 COVID-19 patients with microbiological or radiological evidence of pneumonia. AUC, area under the curve; CI, confidence interval.

|  | Total population |  | Excluding ICU attendance<br>≤72hr of hospital admission |  | Excluding microbiological or<br>radiological pneumonia |  |
| --- | --- | --- | --- | --- | --- | --- |
| Characteristic | AUC | 95% CI | AUC | 95% CI | AUC | 95%CI |
| WCC on admission | 0.79 | 0.72-0.87 | 0.83 | 0.76-0.90 | 0.80 | 0.73-0.88 |
| CRP on admission | 0.65 | 0.56-0.73 | 0.68 | 0.59-0.76 | 0.66 | 0.57-0.74 |
| ΔWCC | 0.77 | 0.70-0.84 | 0.78 | 0.70-0.85 | 0.77 | 0.69-0.84 |
| ΔCRP | 0.76 | 0.68-0.83 | 0.76 | 0.68-0.84 | 0.75 | 0.67-0.83 |
| WCC and ΔWCC | 0.81 | 0.74-0.88 | 0.83 | 0.77-0.90 | 0.81 | 0.74-0.88 |
| WCC and ΔCRP | 0.84 | 0.78-0.91 | 0.87 | 0.81-0.93 | 0.84 | 0.78-0.91 |
| ΔWCC and ΔCRP | 0.80 | 0.73-0.87 | 0.81 | 0.73-0.88 | 0.79 | 0.72-0.86 |
| WCC, ΔWCC<br>and ΔCRP | 0.84 | 0.78-0.91 | 0.87 | 0.80-0.93 | 0.84 | 0.77-0.91 |

Finally, we applied the cut-offs independently derived from the RFH cohort on BH patient data, and observed a similar trade-off between sensitivity and specificity. Using either admission WCC>8.2×10^6^/ml or ΔCRP<0 yielded a CAP sensitivity approaching 95% and negative predictive value of 84% (table 6). Despite the low specificity resulting from these cut-offs, this would still have excluded CAP in 26/56 (46.4%) of COVID-19 patients with no microbiological or radiological evidence of pneumonia and admitted for >48hours. 55 (98%) of these patients were prescribed a 5-day course of antibiotics on admission (203 antibiotic days in total). Stopping antibiotic prescriptions 48 hours into admission in the 26 patients that did not meet either diagnostic criterium for CAP would have saved 51 antibiotic days overall, a 25% reduction in total antibiotic prescriptions in the BH COVID-19 cohort.

**Table 6.** Discriminatory performance of WCC and ΔCRP cut-offs for diagnosis of bacterial pneumonia in BH patients. Populations included were 155 patients admitted to hospital >48hours, including 99 pneumonia cases and 56 COVID-19 cases with no microbiological or radiological evidence of pneumonia.

| Cut-off | Sensitivity % | Specificity % | Positive predictive value % | Negative predictive value % | Positive likelihood ratio | Negative likelihood ratio |
| --- | --- | --- | --- | --- | --- | --- |
| WCC>8.2 | 84.3 | 57.1 | 75.8 | 69.6 | 1.97 | 0.28 |
| WCC>9.5 | 76.4 | 71.4 | 81.0 | 65.6 | 2.67 | 0.33 |
| ΔCRP<3.5 | 66.3 | 73.2 | 79.7 | 57.7 | 2.47 | 0.46 |
| ΔCRP<-15 | 56.2 | 80.4 | 82.0 | 53.6 | 2.86 | 0.55 |
| ΔCRP<0 | 65.1 | 75.0 | 80.6 | 57.5 | 2.61 | 0.46 |
| WCC>8.2 AND ΔCRP<0 | 55.1 | 85.7 | 85.9 | 54.5 | 3.85 | 0.52 |
| WCC>8.2 OR ΔCRP<0 | 94.4 | 46.4 | 73.6 | 83.9 | 1.76 | 0.12 |

## Discussion

Elevated inflammatory responses, high case fatality and bacterial co-infections observed in influenza contribute to frequent antibiotic prescriptions in COVID-19 [2,4,9,27]. However, radiological findings in COVID-19 are heterogenous [28] and microbiological investigations rarely identify pathogenic bacteria [6–9], precluding reliable identification of co-infection. Therefore, novel approaches to exclude bacterial co-infection in COVID-19 are a research priority to facilitate antimicrobial stewardship efforts [14,29,30].

We used patients admitted with CAP prior to the COVID-19 pandemic as a benchmark to define the processes that occur in bacterial pulmonary infections, including bacterial co-infection in COVID-19. This demonstrated that admission WCC (predominantly neutrophils), and CRP can discriminate CAP from both COVID-19 and influenza A/B at a population level. Moreover, WCC and CRP decreased following antibiotic therapy in pneumonia, but not in viral infections. We used these observations to construct a model and decision-making criteria to assist with excluding bacterial co-infection in many cases of COVID-19. We propose that the absence of both admission WCC >8.2×10^6^/ml and a fall in CRP would support stopping antibiotics in almost 50% of COVID-19 patients during their admission, reducing total antibiotic prescriptions in this population by up to 25%. This approach would exceed most antimicrobial stewardship achievements [29,30], and reduce selection for antibiotic resistant bacteria during this pandemic [12,14].

The combination of selected cut-offs yielded the greatest sensitivity for CAP, ensuring ongoing antibiotic treatment where needed, but came at the expense of specificity. Therefore, these criteria remain permissive to excessive antibiotic prescribing in COVID-19, particularly on admission to hospital. PCT can discriminate bacterial from some viral respiratory tract infections [21,22], but has not been systematically compared between bacterial pneumonia and COVID-19. Unfortunately, we could not investigate PCT as it was not measured routinely in our CAP cohorts. Future studies should also assess, alongside admission WCC and ΔCRP, the discriminatory capacity of PCT, D-dimers [9,22,31], and other novel biomarkers, such as transcriptional signatures that quantify inflammatory cytokine activity [32] or those that discriminate bacterial from viral infections [33].

The large, standardised populations studied, use of routinely available clinical investigations, and the reproducibility of our findings in an independent validation cohort population are key strengths of our study. We were also able to convert population level findings into practical diagnostic criteria that can be used in generalised clinical settings. The cut-offs used were derived from a separate population in which they were tested, adding scientific validity to our conclusions, but does not negate individual care providers determining the distribution of admission WCC in their own cases of CAP to define institution-specific IQR cut-offs. In addition, our criteria should not be considered in isolation from clinical decision making. Clinical improvement, reduced supplemental oxygen requirement, and the absence of consolidation on chest radiograph in COVID-19 patients may all contribute to excluding bacterial co-infection and could be used alongside the WCC and ΔCRP criteria to support cessation of antibiotics.

Our study has some notable limitations. First, the populations were identified at non-overlapping times, due to the disproportionate prevalence of COVID-19 cases in 2020. We attempted to mitigate for the enforced use of historical pneumonia and influenza comparator groups by identifying these patients over the same months of 2019 as COVID-19 cases in 2020. We also did not collect clinical severity or outcome data for the patients, and thus we cannot measure a direct impact on prognosis. Second, we used a radiological, but not microbiological, definition of pneumonia, and although standardised, it is possible that some pneumonia cases had non-bacterial aetiology. Third, we inferred that bacterial co-infection in COVID-19 shares pathophysiology and inflammatory marker responses with CAP in the absence of COVID-19. This hypothesis remains untested but is supported by the divergent inflammatory marker responses observed between most COVID-19 and CAP patients. Fourth, we did not include suspected COVID-19 patients with negative SARS-CoV-2 results, therefore our findings may not be applicable to this cohort. Finally, we focused on patient assessments made within 72 hours of admission, and thus our decision-making tools are not applicable to patients discharged before this time or those with prolonged hospital admissions.

In conclusion, we demonstrate that routine clinical parameters, admission WCC and changes in CRP following antibiotic administration, can be translated into a set of diagnostic criteria that can exclude bacterial co-infection in up to half of COVID-19 patients. The routine nature of the investigations required mean that, even in the context of a pandemic, this approach can form the basis of protocols to assist reductions in unnecessary antibiotic prescriptions for viral infections, minimising drug-associated adverse effects and reducing development of antimicrobial resistance.

## Data Availability

Further details on the anonymised clinical data used in this manuscript are available on request from the corresponding author.

## Acknowledgements

We wish to thank members of the RFL Clinical Practice Group for prompt extraction of clinical records.

---

**Figure S1.**
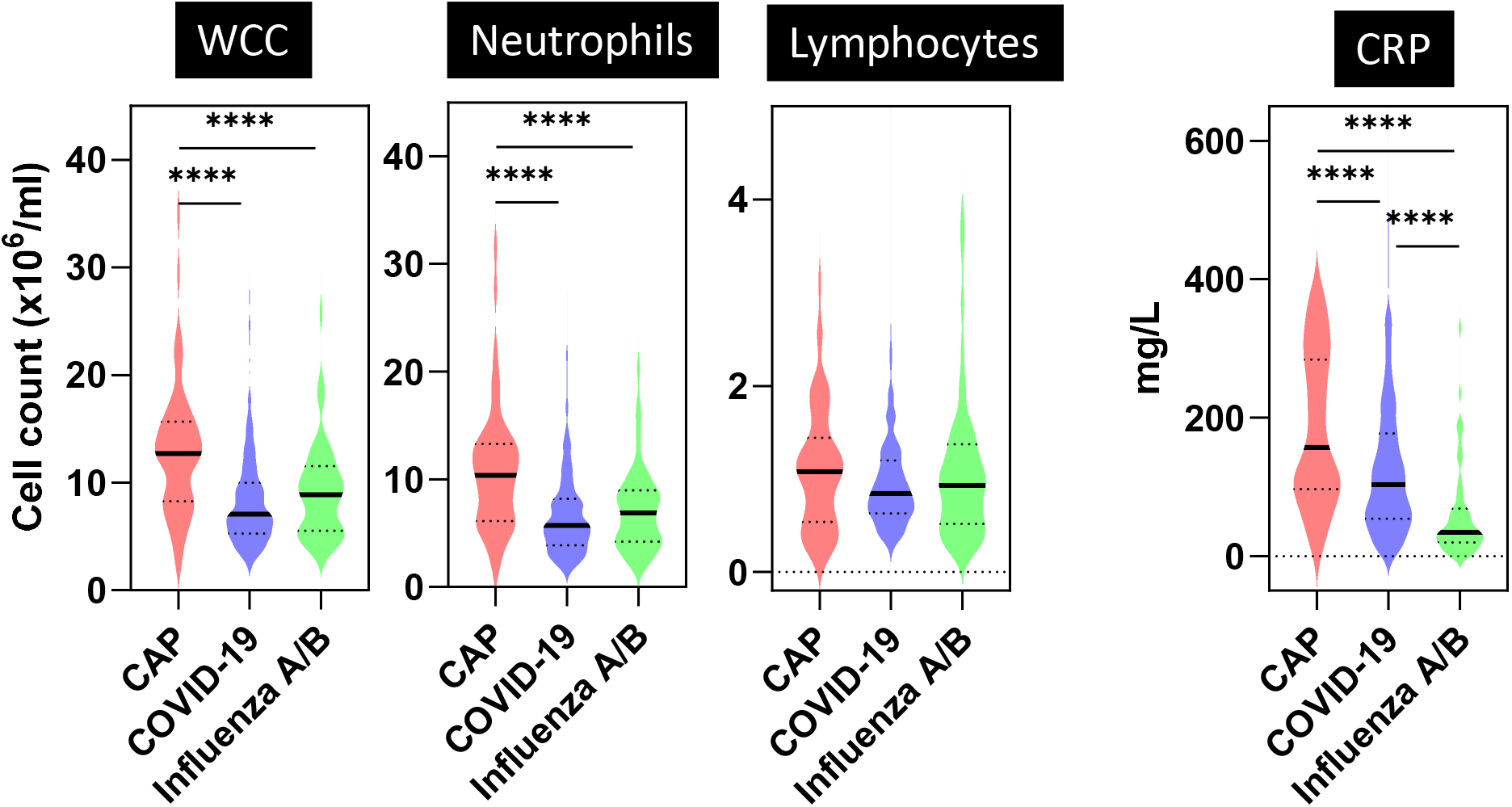
Admission blood samples for RFH patients admitted >48hours. Violin plots represent distribution of values for CAP (n=53), COVID-19 (n=331) and influenza A/B (n=53) patients. Bold lines represent median values. Dotted lines represent IQR values. **** indicates p<0.0001 by Mann-Whitney test.

**Figure S2.**
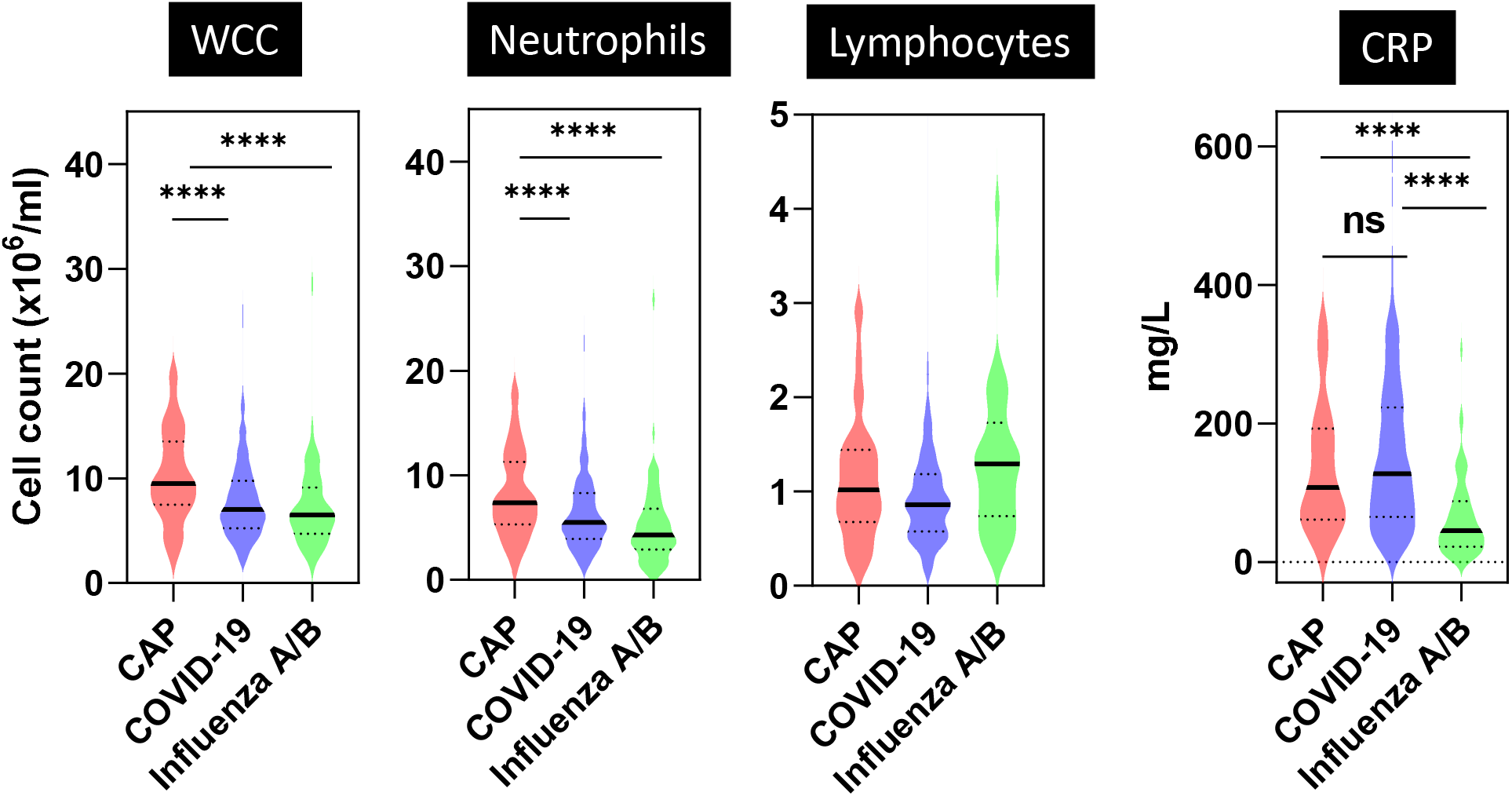
Blood samples collected from RFH patients 48-72 hours into admission. Violin plots represent distribution of values for CAP (n=53), COVID-19 (n=331) and influenza A/B (n=53) patients. Bold lines represent median values. Dotted lines represent IQR values. **** indicates p<0.0001 and ns indicates non-significant by Mann-Whitney test.

**Figure S3.**
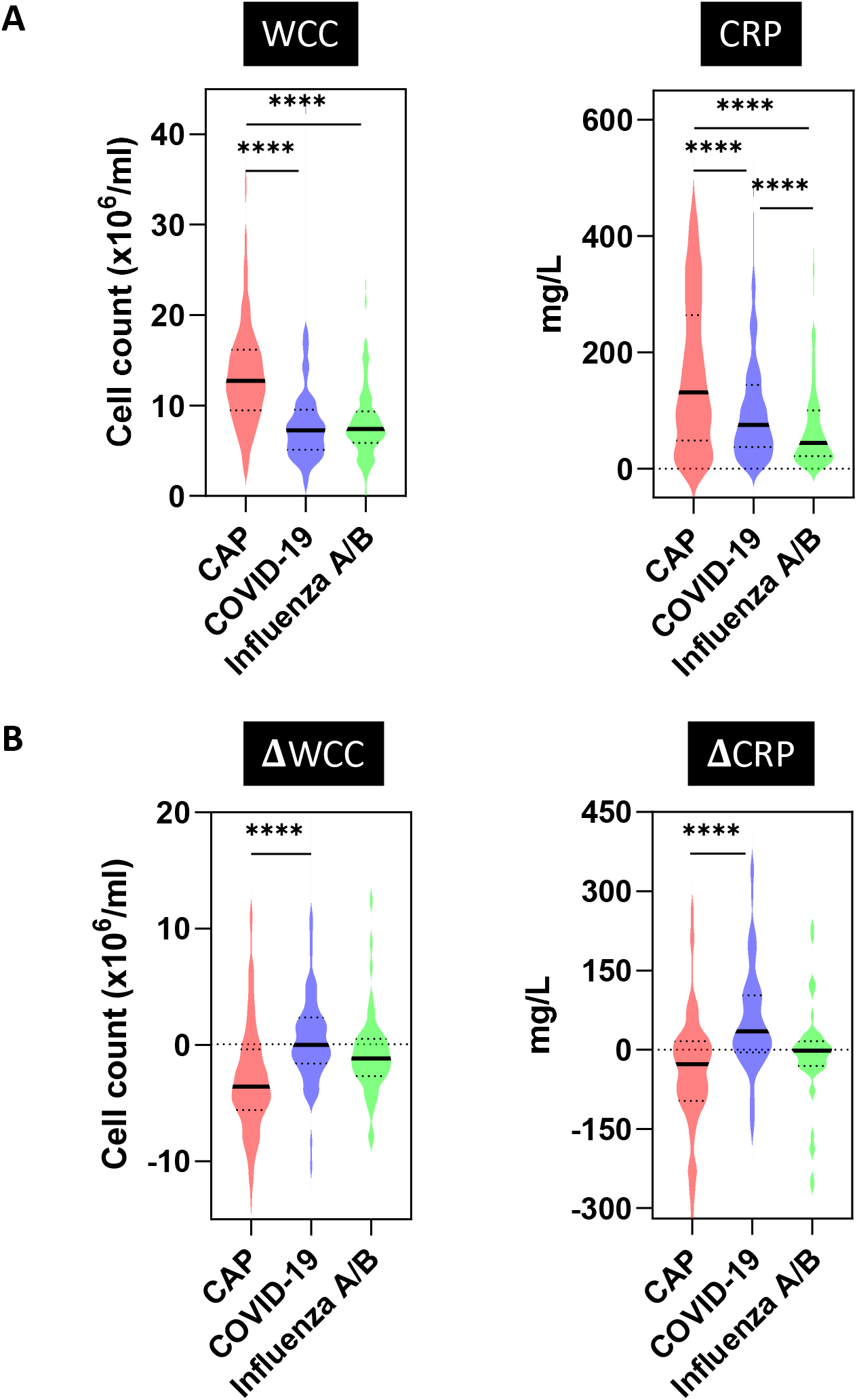
(A) Blood samples collected from BH patients on admission for CAP (n=169), COVID-19 (n=181) and influenza A/B (n=162) patients. (B) Difference (Δ) in blood parameters between samples collected on admission and samples collected 48-72 hours into admission for CAP (n= 99), COVID-19 (n=60) and influenza A/B (n=47) patients. Violin plots represent distribution of values. Bold lines represent median values. Dotted lines represent IQR values. **** indicates p<0.0001 by Mann-Whitney test.

**Table S1.** Bacteria identified by conventional microbiology investigations in RFH and BH patients. Numbers quantity routine clinical samples that yielded specified bacteria from patients described in tables 1 & 4.

|  | Hospital | RFH |  |  | BH |  |  |
| --- | --- | --- | --- | --- | --- | --- | --- |
|  | Disease | CAP | COVID-19 | Influenza | CAP | COVID-19 | Influenza |
| **SPUTUM** | *S pneumoniae* | 2 | 0 | 2 | 0 | 0 | 0 |
|  | *H influenzae* | 2 | 0 | 1 | 1 | 0 | 2 |
|  | *M catarrhalis* | 1 | 1 | 0 | 1 | 0 | 0 |
|  | *S aureus* | 0 | 2 | 1 | 0 | 6 | 2 |
|  | *M pneumoniae* (PCR) | 5 | 2 | 0 | 0 | 0 | 0 |
| **BLOOD** | *S pneumoniae* | 1 | 0 | 1 | 6 | 0 | 1 |
|  | Non-pneumococcal streptococci | 0 | 2 | 0 | 0 | 1 | 0 |
|  | *S aureus* | 0 | 5 | 1 | 0 | 1 | 0 |
|  | Gram negative organisms | 1 | 1 | 2 | 0 | 2 | 0 |
|  | Other | 0 | 4 | 2 | 0 | 1 | 0 |
| **URINE** | *S pneumoniae* antigen | 4 | 3 | 2 | 8 | 0 | 0 |

